# North Carolina macular dystrophy: phenotypic variability and computational analysis of disease-implicated non-coding variants

**DOI:** 10.1101/2021.03.05.21252975

**Authors:** David J. Green, Eva Lenassi, Cerys S. Manning, David McGaughey, Vinod Sharma, Graeme C. Black, Jamie M. Ellingford, Panagiotis I. Sergouniotis

**Affiliations:** Division of Evolution and Genomic Sciences, School of Biological Sciences, Faculty of Biology, Medicines and Health, University of Manchester M13 9PT, UK; Manchester Centre for Genomic Medicine, St Mary’s Hospital, Manchester University NHS Foundation Trust, Manchester M13 9WL, UK; Manchester Royal Eye Hospital, Manchester University NHS Foundation Trust, Manchester M13 9WL, UK; Division of Developmental Biology and Medicine, School of Medical Sciences, Faculty of Biology, Medicines and Health, University of Manchester M13 9PT, UK; Ophthalmic Genetics and Visual Function Branch, National Eye Institute, National Institutes of Health, Bethesda, Maryland, United States

## Abstract

**Purpose:** North Carolina macular dystrophy (NCMD) is an autosomal dominant, congenital disorder affecting the central retina. Here, we report clinical and genetic findings in three families segregating NCMD and use epigenomic datasets from human tissues to gain insights into the effect of NCMD-implicated variants.

**Methods:** Clinical assessment and genetic testing were performed. Publicly-available transcriptomic and epigenomic datasets were analyzed and the ‘Activity-by-Contact’ (ABC) method for scoring enhancer elements and linking them to target genes was used.

**Results:** A previously-described, heterozygous, non-coding variant upstream of the *PRDM13* gene was detected in all six affected study participants (chr6:100,040,987G>C [GRCh37/hg19]). Inter- and intra-familial variability were observed; the visual acuity ranged from 0.0 to 1.6 LogMAR and fundoscopic findings ranged from visually insignificant, confluent, drusen-like macular deposits to coloboma-like macular lesions. Variable degrees of peripheral retinal spots (which were easily detected on widefield retinal imaging) were observed in all study subjects. Notably, a 6-year-old patient developed choroidal neovascularization and required treatment with intravitreal bevacizumab injections. Computational analysis of the five single nucleotide variants that have been implicated in NCMD revealed that these non-coding changes lie within two putative enhancer elements; these elements are predicted to interact with *PRDM13* in the developing human retina. *PRDM13* was found to be expressed in the fetal retina, with highest expression in the amacrine precursor cell population.

**Conclusions:** We provide further evidence supporting the role of *PRDM13* dysregulation in the pathogenesis of NCMD and highlight the utility of widefield retinal imaging in individuals suspected to have this condition.

## INTRODUCTION

North Carolina macular dystrophy (NCMD) is a developmental abnormality affecting the macula, the central part of the retina that is responsible for detailed vision. NCMD is present at birth and rarely progresses. It is inherited as an autosomal dominant trait and although it is thought to be completely penetrant, it exhibits significant intra- and inter-familial variability. The phenotype ranges from visually inconsequential subtle retinal spots to macular coloboma-like lesions associated with significant visual impairment.^1–6^

NCMD was first described in the 1970s in a large pedigree of more than 500 individuals spanning seven generations. This kindred represented a portion of the descendants of two brothers who emigrated from Ireland to the mountains of North Carolina in the early 19^th^ century.^7,8^ Since the first description of the disease, families have been reported in Europe, North America, Asia, and elsewhere in the world.^9–13^ NCMD is genetically heterogeneous but most individuals carry genetic variants in an intragenic region in chromosome 6 located approximately 13,000 base-pairs from the neighboring gene. At least five single nucleotide changes and three copy number variants in this region have been associated with NCMD.^1,3,4,9,14^ These mutations alter DNA sequences that are likely to play a role in regulating the spatio-temporal expression of a retinal transcription factor *PRDM13. PRDM13* is expressed in the fetal retina but is not found in adult tissues.^1^ Over-expression of this gene has been shown to affect retinal development in non-primate animal models.^15–17^ Intriguingly, changes near *PRDM13* that have been associated with NCMD have also been implicated in a different condition, progressive bifocal chorioretinal atrophy (PBCRA). Although both these disorders affect the macula from birth, the latter is typically associated with disease progression and electrophysiological evidence of widespread retinal dysfunction.^2^ It is also noteworthy that a number of individuals with a phenotype indistinguishable from NCMD have been found to have copy number variants in a different locus in chromosome 5.^1,2^

NCMD is part of an expanding group of Mendelian conditions caused by non-coding genetic variants that have an effect on gene expression (e.g., aniridia,^18^ limb malformations^19^). These variants often alter the sequence of regulatory elements, including transcriptional promoters and enhancers. Enhancers are short stretches of DNA, typically a few hundred base-pairs long, that bind transcription factors and enhance the expression of specific genes.^20^ There are hundreds of thousands of enhancers in the human genome but our understanding of their properties and repertoire is limited (e.g., where they reside, what genes they mediate their effects through, in which cells and in which specific time of development they act). Notably, our ability to detect and annotate enhancer elements is being transformed by emerging resources and tools such as the Encyclopedia of DNA Elements (ENCODE) 3 dataset,^21^ the Developmental Single Cell Atlas of Gene Regulation and Expression (DESCARTES),^22,23^ and the Activity-by-Contact (ABC) model.^24^ The ENCODE project consortium and the DESCARTES team have generated extensive functional genomic datasets across many cell and tissue contexts. Experiments conducted include assays aiming to identify enhancer elements such as DNase-seq (DNase I hypersensitive site sequencing), H3K27ac ChIP-seq (chromatin immunoprecipitation sequencing mapping H3K27 acetylation signals), and ATAC-seq (assay for transposase-accessible chromatin using sequencing), as well as assays studying the 3-dimensional architecture of the genome such as Hi-C (all-versus-all chromosome conformation capture).^25^ ABC in contrast is a method that uses these epigenomic data to predict which enhancers regulate which genes. This model is based on the notion that an enhancer’s effect on a gene depends on the enhancer’s strength (estimated using chromatin accessibility data including ATAC-seq and H3K27ac ChiP-seq) weighted by how often it comes into 3-dimensional contact with the gene promoter (estimated using Hi-C data).^24^ Although this method has been shown to be accurate, a limiting factor is the availability of tissue-specific chromatin accessibility datasets. Comprehensive tissue-specific Hi-C data are also lacking but the developers of ABC expect that Hi-C profiles averaged across a selection of cell types are adequate to map enhancer-gene connections. However, it remains possible that tissue- and context-specific data are required in some cases as the 3-dimensional conformation of the genome shows some plasticity during development.^26^ Although epigenomic datasets from various eye tissues are becoming increasingly available,^27^ there have been no systematic efforts to characterize enhancers that contribute to human vision.

In this report, we discuss phenotypic variability in individuals who have NCMD and are heterozygous for a specific non-coding variant, chr6:100,040,987G>C (GRCh37/hg19). We then utilize the ABC method and human transcriptomic and epigenomic datasets to gain mechanistic insights into the role of non-coding variation in NCMD.

## METHODS

### Participant ascertainment, phenotypic data collection & clinical genetic testing

Six individuals with a diagnosis of NCMD were retrospectively ascertained through the database of the North West Genomic Laboratory Hub, Manchester, UK. The study participants originated from three reportedly unrelated families who had European ancestries. The genetic and, to an extent, the clinical findings from two of the three affected individuals in one of these families have been previously reported.^14^ Ethics committee approval for the study was obtained from the North West Research Ethics Committee (11/NW/0421 and 15/YH/0365) and all investigations were conducted in accordance to the tenets of the Declaration of Helsinki.

All study participants were diagnosed with NCMD through the tertiary ophthalmic genetics service at Manchester University NHS Foundation Trust, Manchester, UK. Clinical assessment included visual acuity testing, dilated fundus examination, digital widefield fundus imaging, fundus autofluorescence (FAF) imaging, optical coherence tomography (OCT), and OCT-angiography (OCT-A). The Optos system (Optos PLC, Dunfermline, Scotland, UK) was used to obtain widefield images, the Topcon DRI OCT Triton device (Topcon GB, Newberry, Berkshire, UK) was used to obtain OCT and OCT-A scans, and the Spectralis system (Heidelberg Engineering, Heidelberg, Germany) was used to acquire FAF and OCT images in study participants.

Genetic testing was performed at the North West Genomic Laboratory Hub Diagnostic Laboratory, a UK Accreditation Service Clinical Pathology Accredited medical laboratory (Clinical Pathology Accredited identifier, no. 4015). DNA samples from all study subjects were screened using Sanger sequencing (BigDye v3.1) for the presence of known NCMD-implicated variants in chromosome 6 (upstream of the PRDM13 gene^1^). Two affected individuals also underwent genome sequencing as previously described (case 89794.1^28^ and case 75898.2^14^, see Figure 1 & Table 1); the generated data were used for haplotype analysis (Supplementary Table 1).

**Table 1.** Clinical characteristics of individuals carrying the chr6:100,040,987G>C (GRCh37/hg19) change in heterozygous state

| patient | gender | age<br>(at presentation;<br>age at last<br>examination) | LogMAR vision<br>at last<br>examination<br>(right; left) | main fundoscopic<br>findings |
| --- | --- | --- | --- | --- |
| 75898.1 | male | 3; 7 | 0.5; 1.6 | coloboma-like macular lesions; subfoveal scarring and fluid due to neovascularization; subtle peripheral white spots |
| 75898.2 | female | 4; 36 | 0.5; 0.5 | coloboma-like macular lesions with associated macular white spots and a degree of foveal sparing; peripheral white spots |
| 75898.3 | female | 4; 58 | 1.5;1.5 | coloboma-like macular lesions with associated macular white spots; peripheral white spots |
| 123660.1 | female | 35; 37 | 0.0; 0.0 | macular and peripheral white spots |
| 89794.1 | male | 44; 50 | 0.0; 0.0 | macular and peripheral white spots |
| 89794.2 | male | 46; 52 | 0.0; 0.0 | macular and peripheral white spots |
| 89794.1 and 89794.2 are brothers; 75898.2 is the daughter of 75898.3 and mother of 75898.1. The genetic and phenotypic findings in 75898.2 and 75898.3 are also discussed in |  |  |  |  |

**Figure 1.**
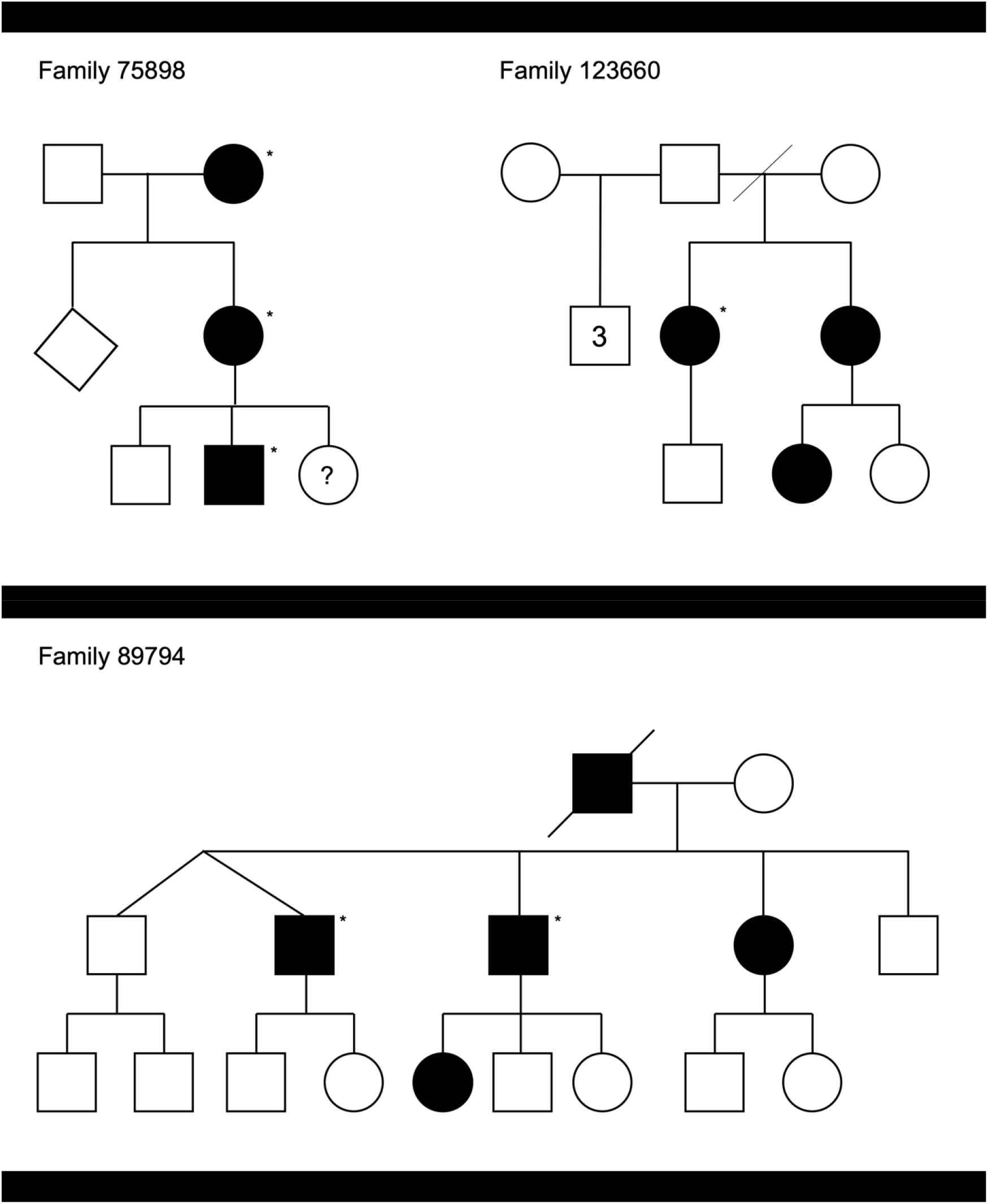
Pedigrees from three families segregating North Carolina macular dystrophy. Study participants are highlighted with asterisks. We note that findings from the two affected females from family 75898 are also discussed in ^14^

### Analysis of variants implicated in North Carolina macular dystrophy

To gain insights into the frequency of NCMD-implicated variants in the general population, we queried the Genome Aggregation Database (gnomAD; version 3.1) and the NHLBI Trans-Omics for Precision Medicine variant database (TOPMed; freeze 8). Neither of these cohorts is known to be enriched for patients with ophthalmic conditions.^29,30^

All variants known to be associated with NCMD (including the chr6:100,040,987G>C change detected in the study participants) were assessed using *in silico* tools that attempt to predict the deleteriousness of non-coding sequence alterations. The following tools were utilized: CADD v1.6,^31^ nCER v2,^32^ and regBase v1.1.^32^ Position-specific evolutionary constraint was evaluated using GERP (Genomic Evolutionary Rate Profiling) scores obtained through the UCSC (University of California, Santa Cruz) browser.^33,34^

To investigate the functional effects of the variants implicated in NCMD, we queried the v109 release of the ENCODE portal for putative enhancer elements spanning the location of each variant; the SCREEN (Search Candidate cis-Regulatory Elements by ENCODE) v10 interface was used.^21^ We also inspected the eQTL catalogue and the GTEx (Genotype-Tissue Expression) and EyeGEx (Eye Genotype Expression) datasets for expression quantitative trait loci (eQTL) overlapping any of the detected elements (in addition to elements predicted in our subsequent analyses).^35,36^ We then queried the DESCARTES chromatin accessibility database for the location of NCMD-implicated variants across all available tissues.

The ABC method for scoring enhancer elements and linking them to target genes was subsequently utilised.^24^ Raw ATAC-seq and H3K27ac ChIP-seq reads from macular and retinal tissue were obtained from a publicly-available dataset (GEO accession: GSE137311).^27^ The data were then processed using a consistent pipeline. The raw reads were trimmed for quality and adaptor content using the fastp tool with default quality control options.^37^ The trimmed ATAC-seq and ChIP-seq reads were aligned to the GRCh37/hg19 reference sequence using bowtie2 version 2.3.0 with the default parameters.^38^ The alignments were inspected in picard tools version 2.1.0 (https://broadinstitute.github.io/picard/) and duplicate reads were removed. Only reads with a mapping quality ≥ 30 were retained in subsequent analyses after filtration using the samtools “-view” command (version 1.9).^39^

The ABC method was then used to produce a set of potential enhancers using default parameters. As described in Fulco *et al*., initial peaks were obtained utilizing the MACS2 (Model-based Analysis of ChIP-Seq 2) peak caller on aligned ATAC-seq reads with a lenient p-value using the following options “-g hs, -p .1, --call-summits”.^40^ The peaks were then extended by 250 base-pairs from their summits to produce a set of 500 base-pair “candidate regions”; overlapping regions were then merged. This resulted in a broad set of potential regulatory elements whose activity could be scored. To accomplish this, the ATAC-seq and H3K27ac ChIP-seq reads overlapping each candidate region were counted and averaged over biological replicates using the ABC “call neighborhoods” script. Finally, Hi-C data from human embryonic stem cells were downloaded from the juicebox archive at 5,000 base-pair resolution.^26^ Averaged Hi-C data provided by the ABC GitHub page were also used; these data were produced by averaging the Hi-C profiles from 10 cell types (GM12878, NHEK, HMEC, RPE1, THP1, IMR90, HUVEC, HCT116, K562, and KBM7). ABC scores were calculated by combining the estimated contact frequency (between the candidate region and the promoter of a gene) with the observed activity using the default parameters.

### Analysis of PRDM13 gene expression in the human retina

To determine the expression level of *PRDM13*, we utilized a resource collecting human RNA-seq datasets: eyeIntegration.^41^ All retinal tissue subtypes, RPE subtypes, and progenitor subtypes were queried. As eyeIntegration does not contain data on macula, *PRDM13* expression was also estimated from three macula RNA-seq samples included in the GEO dataset GSE137311.^27^ In short, the raw reads were quantified using salmon^42^ (according to the same protocol used to build the eyeIntegration dataset) and the value in TPM (transcripts per million) was calculated using tximport to merge reads to the gene level with the “lengthScaledTPM” option. The source code for this pipeline can be found at: www.githubdavemcg.com/EiaDBuild/. DESCARTES (https://descartes.brotmanbaty.org/) was also queried for *PRDM13* expression. Finally, *PRDM13* expression was determined in individual cell types using the PLatform for Analysis of scEiad (plae v0.43) database (https://plae.nei.nih.gov/).

## RESULTS

### Clinical characteristics of study participants

All six study participants were heterozygous for the chr6:100,040,987G>C variant. This change has been previously detected in a number of families segregating NCMD^1,3,4,14^ and appeared to be on the same genetic background (haplotype) in at least two of the families included in this study (families 75898 and 89794; Supplementary Table 1).

The age at presentation ranged from 3 to 46 years of age and the visual acuity ranged from 0.0 to 1.6 LogMAR. Three related patients presented in the first years of life and had vision of 0.5 LogMAR or worse, while the remaining three patients had normal vision and were only found to have drusen-like macular lesions on routine eye tests. The clinical findings are discussed in Table 1, the pedigrees are shown in Figure 1, and fundus imaging results are shown in Figures 2 and 3. The central macular changes resembled fine, confluent drusen; these lesions were hyperautofluorescent on FAF imaging and only partially corresponded to visible OCT changes. A notable finding in all participants was that of characteristic yellow-white drusen-like retinal lesions in the far periphery; these generally had a linear, radial configuration and could be detected on widefield imaging (Figure 3). The youngest study participant developed choroidal neovascularization in both maculae (diagnosed at 6 years of age). An active lesion was noted in the right eye on OCT-A and treatment with intravitreal bevacizumab injections was initiated. However, persistent subretinal fluid was present after four injections.

**Figure 2.**
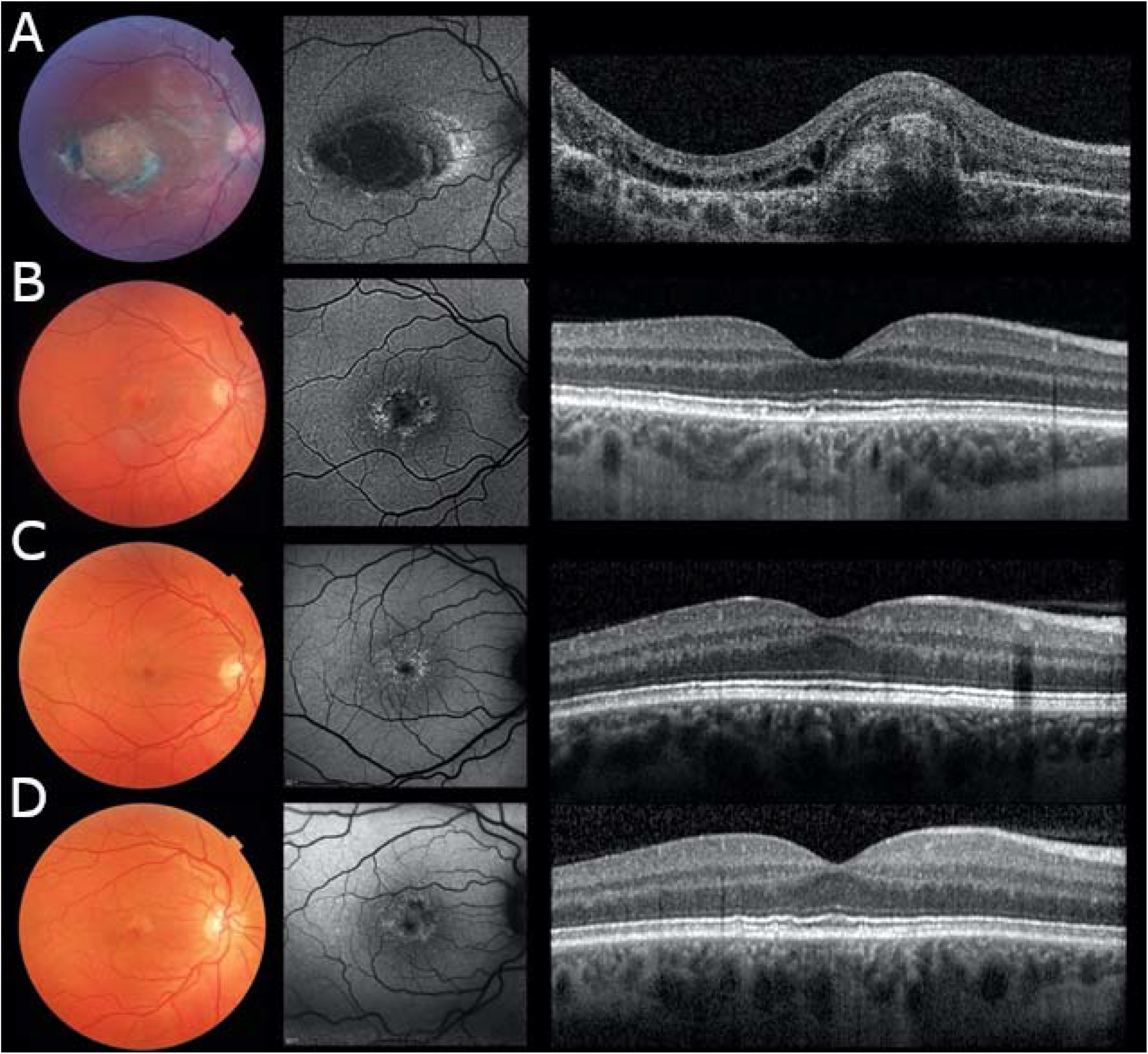
Color fundus photography, fundus autofluorescence imaging and OCT from four individuals who have North Carolina macular dystrophy and are heterozygous for the chr6:100,040,987 variant (GRCh37/hg19). [A] Macular findings in a 6-year-old child are shown. A macular coloboma-like excavation is seen; there is extensive subretinal fibrosis centrally with residual pockets of intraretinal fluid despite treatment with four intravitreal bevacizumab injections. Images of the affected mother and grandmother of this proband can be found in Ellingford et al. (2017).^14^ [B] Macular findings in a 36-year-old proband are shown. Subtle confluent yellow-white specks are noted in the central macula. These are more visible on fundus autofluorescence imaging and correspond to hyperautofluorescent lesions. Only a subset of these changes were readily identifiable by OCT. No inner retinal layer abnormality could be detected. [C,D] Macular findings in a 44-year-old proband and his 48-year-old brother are shown. Yellow-white foveal lesions, similar to those observed in the 36-year-old individual discussed above, are noted. There was a high degree of interocular symmetry and only data from the right eye are shown.

**Figure 3.**
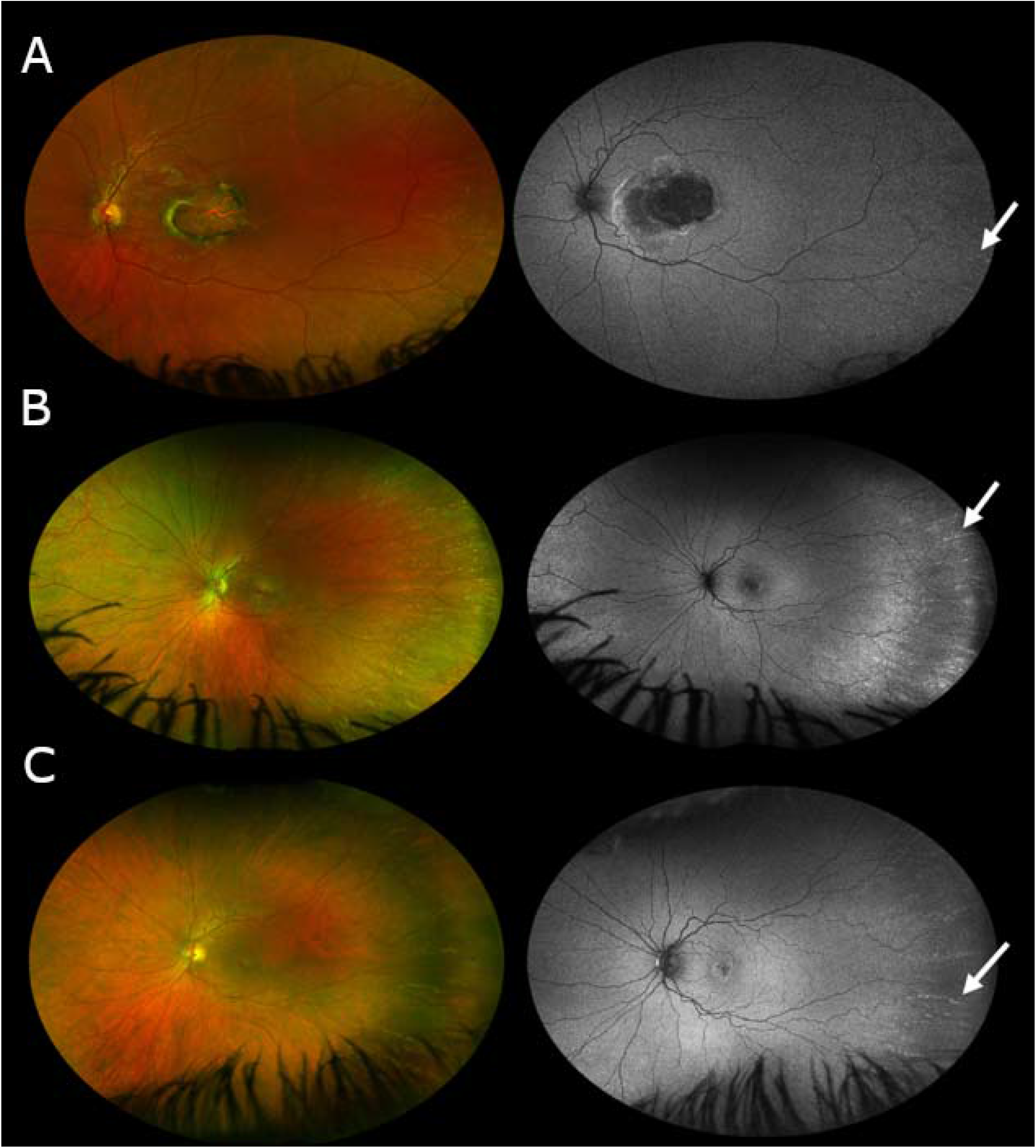
Digital widefield fundus imaging and fundus autofluorescence imaging in three probands who have North Carolina macular dystrophy and are heterozygous for the chr6:100,040,987 variant (GRCh37/hg19). Images from a 6-year-old [A], a 36-year-old [B], and a 48-year-old [C] are shown. Yellow-white retinal spots, often very prominent and in radial alignment, are noted in the far temporal periphery of these three individuals. These spots correspond to hyperautofluorescent lesions on fundus autofluorescence imaging. White arrows are used to highlight some of these changes. Inferior lash artefact is noted in some images. There was a high degree of interocular symmetry and only data from the left eye are shown. It is worth noting that although there was significant phenotypic variability, widefield imaging revealed a maculopathy combined with peripheral drusen-like spots in all study participants.

### In silico analysis of variants associated with North Carolina macular dystrophy

To date, five NCMD-implicated single nucleotide variants have been reported in the biomedical literature. These variants were found to be extremely rare upon inspection of large-scale genomic datasets (gnomAD and TOPMed; Table 2). Notably, the chr6:100,040,987G>C change that was detected in the participants of this study was only identified in a single individual in gnomAD; this person was estimated to have European ancestry. None of the five NCMD-implicated variants was predicted to be deleterious by CADD. According to nCER, the variants ranged from low to middle percentiles.^43^ The regBase Phred-like scores ranged from 5.7 to 19.8 on a Phred-like scale from 1 to 99, with higher scores suggesting greater pathogenicity. Inspection of the UniPROBE (Universal PBM Resource for Oligonucleotide Binding Evaluation) database^44^ revealed that none of these disease-causing variants is expected to significantly impact on a transcription factor binding site. The GERP scores ranged from -2.44 to 2.52, suggesting variable evolutionary constraint (Table 2).

**Table 2.** Single nucleotide variants in chromosome 6 associated with North Carolina macular dystrophy.

| Variant (GRCh37/hg19) | Variant (GRCh38/hg38) | gnomAD allele count | TOPMed allele count | CADD-Phred score | regBase score | ncER percentile | GERP score | number of families (number of affected) | reference |
| --- | --- | --- | --- | --- | --- | --- | --- | --- | --- |
| chr6:100,040,906G>T | chr6:99,593,030G>T | 0/~152,100 | 0/~264,690 | 1.672 | 19.8 | 81.33 | -0.01 | 6 (65) | <sup>1</sup> |
| chr6:100,040,974A>C | chr6:99,593,098A>C | 0/~152,100 | 0/~264,690 | 1.465 | 8.3 | 73.36 | -2.19 | 1 (6) | <sup>9</sup> |
| chr6:100,040,987G>C | chr6:99,593,111G>C | 1/152,188 | 0/~264,690 | 9.133 | 19.5 | 79.83 | 2.52 | 8 (20) | <sup>1,3,4,14</sup> |
| chr6:100,041,040C>T | chr6:99,593,164C>T | 0/~151,800 | 0/~264,690 | 5.647 | 16.9 | 69.37 | 1.72 | 1 (2) | <sup>1</sup> |
| chr6:100,046,783A>C* | chr6:99,598,907A>C | 0/~152,100 | 0/~264,690 | 2.528 | 5.7 | 15.60 | -2.44 | 1 (1) | <sup>2</sup> |

gnomAD, Genome Aggregation Database v3.1 dataset; GERP, Genomic Evolutionary Rate Profiling; TOPMed, NHLBI Trans-Omics for Precision Medicine variant database freeze 8; CADD, combined annotation dependent depletion; ncER, non-coding essential regulation.
The CADD score ranges from 1 to 99; a higher score indicates greater pathogenicity. Values $\geq 10$ are predicted to be the 10% most deleterious substitutions and $\geq 20$ in the 1% most deleterious.
The ncER scores are percentiles. The higher the percentile, the more likely that a region is vital in terms of regulation. There is no agreed cut-off for deleteriousness but the original report focused on assessing the predictive power of the higher percentiles (95<sup>th</sup> and above).<sup>43</sup>
The regBase scores are Phred-like and calculated using the “PAT” model, which attempts to determine whether a variant is deleterious. The scores range from 1 to 99; a higher score indicates greater pathogenicity.<sup>32</sup>
The GERP scores represent position-specific evolutionary constraint. Positive values suggest evolutionarily constrained positions, with a cut-off of 2 providing high sensitivity.<sup>33</sup>
\*this variant has also been associated with progressive bifocal chorioretinal atrophy (PBCRA). Notably, a nearby variant, chr6:100,046,804T>C (GRCh37/hg19), has been found to cause PBCRA in six affected individuals from two families (see also Figure 4).<sup>2</sup>

The degree to which NCMD-implicated variants map to putative enhancer elements was then investigated. Two such elements were found in the ENCODE dataset to span the locations of all five disease-associated variants (accessions: EH37E1264999 and EH37E1265001l; DNase Z-scores: 3.81 and 1.70). We used the ABC method to assign these enhancers to target genes. Publicly-available chromatin accessibility (ATAC-seq and H3K27ac ChIP-seq) datasets from macula and retina were utilized.^27^ The Hi-C data recommended and provided by Fulco *et al*.^24^ were also used; these consist of a Hi-C profile that was averaged over a number of cell types. With this input, ABC did not yield any predictions for the regions spanning the location of NCMD-implicated variants. We then used the same ATAC-seq and H3K27ac ChIP-seq data but this time combined with Hi-C data from human embryonic stem cells. In macula, two predicted enhancers encompassed the locations of all five variants, and these regions were linked to the *FLH5, SIM1*, and *PRDM13* genes.

As previous reports have associated *PRDM13* with NCMD,^2,8^ we then looked for enhancer elements predicted to be linked to *PRDM13*. Only elements that were within 100,000 base-pairs of the transcription initiation site were analyzed. This yielded a total of 53 candidate regions in macular tissue and 47 candidate regions in retinal tissue. To determine which of these candidates have the strongest regulatory effects on *PRDM13*, putative enhancers were ranked according to the overall ABC score and to the individual components of this score (i.e. activity and contact) in isolation. The top 10 predicted elements ranked by their ABC scores are shown in Supplementary Tables 2 and 3.

One previously reported NCMD-associated variant, chr6:100,046,783A>C, fell within a predicted enhancer that spanned approximately the same coordinates in both macula and retina (chr6:100,046,330-100,046,830 and chr6;100,046,348-100,046,848, respectively) (Figure 4). In terms of ranking, this enhancer had low scores for activity but relatively high scores for Hi-C contact when data from embryonic stem cells were used. Another four variants (chr6:100,040,906G>T, chr6:100,040,974A>C, chr6:100,040,987G>C, and chr6:100,041,040C>T) fell within a predicted enhancer spanning the chr6:100,040,653-100,041,153 region. This enhancer was found in macular tissue but not in retina, and also had low scores for activity but relatively high scores for Hi-C contact.

**Figure 4.**
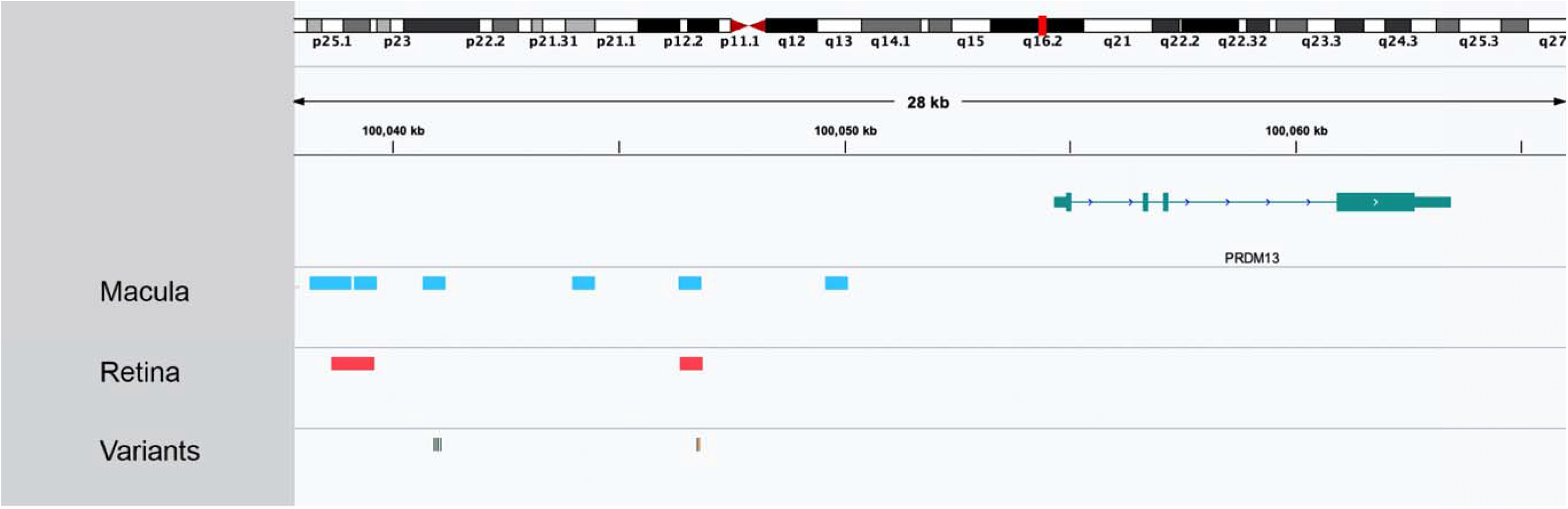
Location of enhancer elements upstream of *PRDM13* in macula and retina as predicted by the Activity-by-Contact (ABC) model.^24^ A putative enhancer encompasses the chr6:100,040,987G>C (GRCh37/hg19) variant, which was carried by the six study participants (green line in the variant row of the Figure). The enhancer encompassing this variant was not detected in retinal tissue and was only highlighted when a dataset from macular tissue was utilized. This enhancer encompasses another four variants that were previously identified in families with North Carolina macular dystrophy (NCMD): chr6:100,040,906G>T, chr6:100,040,974A>C, chr6:100,040,987G>C, and chr6:100,041,040C>T. A different candidate enhancer that was predicted to be functional in both macular and retinal tissue was altered by the chr6:100,046,783A>C variant. This change has been identified in a single family with two affected individuals, a mother and her child; while the mother had typical findings of NCMD, the child had broader retinal involvement in keeping with a related developmental disorder, progressive bifocal chorioretinal atrophy (PBCRA).^2^ Notably, an adjacent variant, chr6:100,046,804T>C (orange line in the variant row of the Figure), has been reported to cause PBCRA in six affected individuals from two families.^2^ The fact that the candidate enhancer encompassing these two PBCRA-implicated variants appears to have a role in both macular and retinal tissue may explain why individuals carrying these changes often develop a phenotype that is more severe than NCMD and is associated with generalized retinal dysfunction.

Finally, we could not find any significant single-tissue eQTLs within any of the enhancer elements found in ENCODE or those predicted by the ABC method. Data from multiple tissues were inspected (including retinal samples included in EyeGEx).

### PRDM13 gene expression in the developing and adult human retina

To gain insights into the cellular context in which the *PRDM13* gene product exerts its function, we evaluated the levels of gene expression in a number of relevant tissues (Figure 5). The expression levels were significantly higher in the retina and macula than in the RPE, with the highest level found in fetal retinal tissue. Analysis of single-cell RNA-seq datasets revealed that *PRDM13* is expressed in progenitor cells (including amacrine/horizontal precursors), with higher expression in the amacrine population. *PRDM13* had low expression levels (<0.2 TPM) in all tissues found in the GTEx project dataset. For further detail on the expression of *PRDM13* across tissues and cell types, please see Supplementary Table 4 and Supplementary Figure 1.

**Figure 5.**
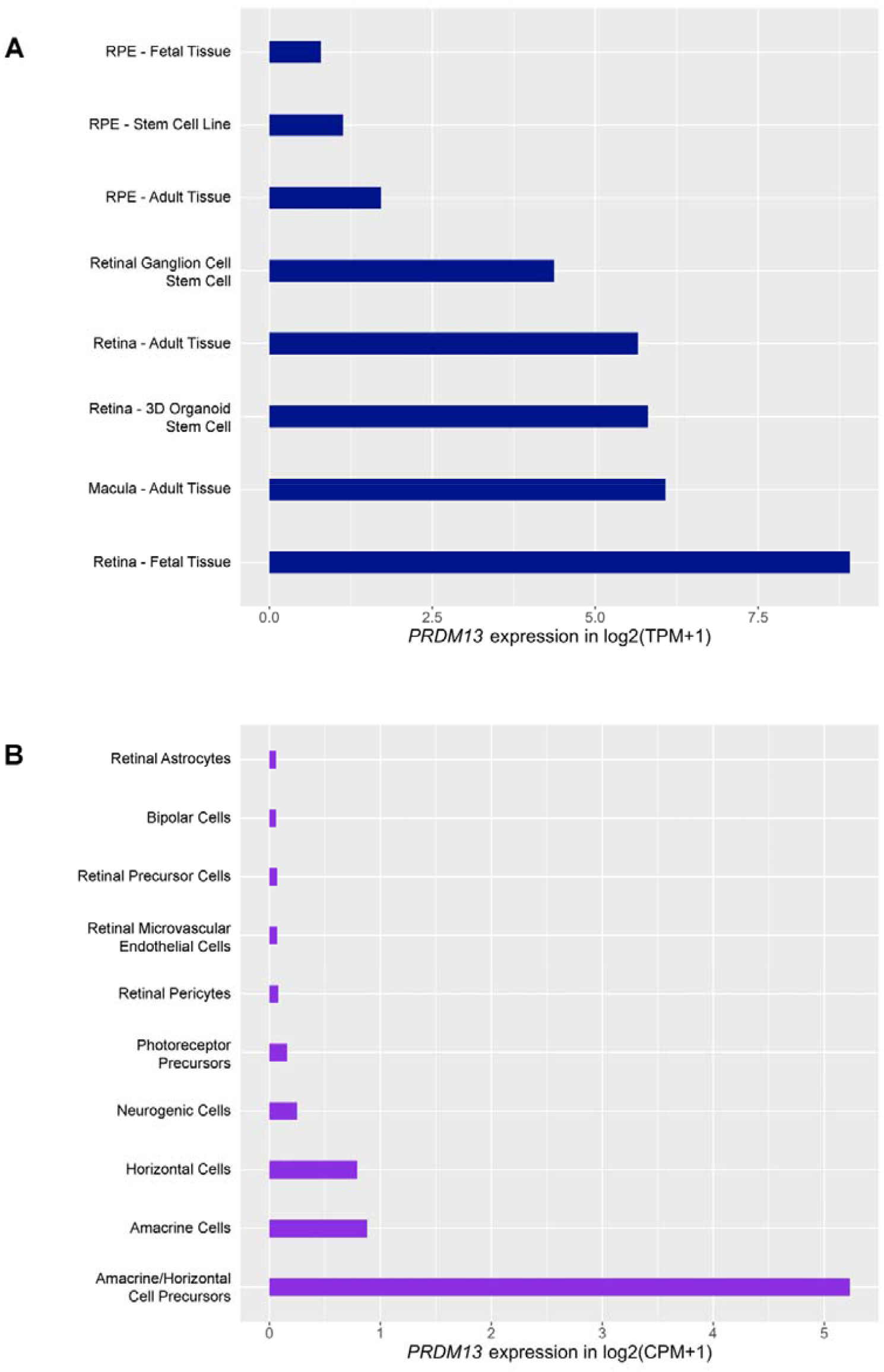
Expression levels of *PRDM13* in different human tissues and cells. [A] gene expression of *PRDM13* in human tissues from bulk RNA, taken from eyeIntegration (https://eyeintegration.nei.nih.gov/:v1.05). [B] gene expression of *PRDM13* in cell populations from single-cell RNA-seq studies (https://plae.nei.nih.gov). It is noteworthy that PRDM13 had very low expression levels (<0.2 TPM) in all extraocular tissues included in the Genotype-Tissue Expression (GTEx) project dataset.^35^ TPM, transcripts per million. CPM, counts per million. RPE, retinal pigment epithelium.

## DISCUSSION

In this study, we report clinical and genetic findings from six patients with NCMD. We highlight the utility of widefield retinal imaging in individuals suspected to have NCMD (Figure 3) and discuss the need for monitoring for choroidal neovascularization, particularly in young affected children. Furthermore, we perform computational analysis of NCMD-associated changes, with the results further supporting the role of *PRDM13* dysregulation in the pathogenesis of the condition.

Phenotypic variability is incompletely understood but common in NCMD.^1,3^ Thus, it is not surprising that significant differences in clinical presentation were observed among the participants of this study. It is worth highlighting though that the three mildest cases were asymptomatic and were initially incorrectly diagnosed in adulthood as having a form of early-onset macular drusen; OCT imaging revealed that these macular changes were unlike typical drusen (Figure 2) and genetic testing highlighted the congenital nature of these lesions. It is noteworthy that histopathologic examination of an enucleated eye from a 72-year-old patient with a clinical diagnosis of NCMD revealed sub-RPE deposits.^45^ However, OCT findings from this and other studies^9^ suggest that at least a subset of these lesions are anterior to the RPE. The most severely affected participant in this study was a 6-year-old male patient who had coloboma-like excavation and subretinal fibrosis in both maculae. Notably, he experienced moderate/severe visual loss due to the formation of choroidal neovascularization. This is a rare but sight-threatening complication of NCMD that can occur in both children and adults.^1,5,46,47^ A previous report has shown that intravitreal bevacizumab injections can improve vision and decrease intraretinal fluid. However, multiple injections (in a treat-and-extend protocol) may be required.^46^

We highlight an under-recognized fundoscopic finding of NCMD: multiple drusen-like spots in the peripheral retina. These were noted in all study participants and were easily detected on widefield retinal imaging (Figure 3). Although this feature is not pathognomonic, it prompted targeted genetic testing in one of the probands. Similar retinal abnormalities have been previously described in a number of affected families, including in the original NCMD kindred.^5,6,11,48,49^ These peripheral lesions should therefore be sought after in all individuals with a clinical presentation suggestive of NCMD and their presence should provide additional justification for requesting focused genetic screening. Further retinal imaging, including widefield OCT, is expected to provide important insights into the nature of this common but frequently overlooked feature of NCMD.

Although a number of non-coding genetic variants have been associated with NCMD, our understanding of the molecular pathology of this condition remains incomplete. The recent emergence of comprehensive functional genomic datasets from adult and fetal human tissues^21,22,27^ has enabled in-depth analysis of non-coding changes like the ones implicated in this disorder. We found that all NCMD-causing variants fall within enhancer elements that appear to be active during development and to be linked to the retinal transcription factor *PRDM13*. A number of observations relating to our *in silico* analysis are worthy of further discussion. First, the candidate enhancer that encompasses chr6:100,040,987G>C, the variant carried by the six study participants, was absent when retina-derived epigenomic data were used and was only highlighted when a dataset from macular tissue was utilized (Figure 4). Second, there were stark differences in the computational predictions of the ABC algorithm depending on the type of Hi-C data used as input (averaged/non-tissue-specific *vs*. embryonic tissue-specific profiles). This is notable as the developers of the ABC algorithm concluded that a Hi-C profile generated by averaging over several tissue types performed equally well to tissue-specific data. However, our findings indicate a potential pitfall in using averaged Hi-C data for genes that are most highly expressed during development.

The human retina emerges in three main stages. The early retina is characterized by retinal progenitor proliferation and RPE emergence (5–7 post-conception week); this is followed by ganglion cell production and initiation of the programs that underlie the development of horizontal cells, amacrine cells, and cone photoreceptors (7–10 post-conception week). Subsequently, cone, amacrine, rod, bipolar, and Muller glia cells sequentially emerge (12–18 post-conception week).^50,51^ Interestingly, the morphological differentiation of the fovea is completed earlier than other regions for all cell types.^50^ Our gene expression analyses of human tissues revealed that *PRDM13* is predominantly expressed in the retinal progenitor and the amacrine cell populations. Amacrine cells are the most diverse class of retinal neurons and are often subdivided based on the expression of inhibitory γ-aminobutyric (GABA) or glycine. Studies in animal models have shown that *PRDM13* is a key determinant of amacrine cell fate and promotes the generation of amacrine cell subtypes (with a bias towards a glycinergic phenotype). Given that duplications of the *PRDM13* region are an established cause of NCMD, it has been proposed that over-expression of this gene during development is the main disease mechanism.^15^ This is supported by findings in Drosophila, Xenopus, and murine retinae, although none of these animal models have a macula.^16,17^ How impaired amacrine cell specification leads to macular abnormalities remains unclear. It is noteworthy though that the inner retinal layers (including the amacrine cell bodies) appeared normal on OCT imaging (Figure 2). Previous studies have however reported cases with subtle foveal hypoplasia^3^ or electrophysiological findings^15^ suggesting amacrine cell abnormalities. These are however sporadic reports and in-depth phenotyping of more affected individuals will provide further insights.

In conclusion, we have used human transcriptomic and epigenomic datasets to study the disease mechanism of NCMD. We highlight the value of computational approaches for the evaluation of candidate non-coding variants and discuss the importance of taking spatio-temporal context into account in these analyses. Furthermore, we report a common peripheral retinal finding challenging the notion that NCMD is a disorder strictly confined to the macula.

## Data Availability

All data are publicly available. Epigenomic and transcriptomic data are available through GEO (accession: GSE137311). The ABC enhancer prediction tool is available on GitHub (https://github.com/broadinstitute/ABC-Enhancer-Gene-Prediction).

## Acknowledgements

We acknowledge the following sources of support and funding: Christopher Green Doctoral Fellowship (DJG), UK National Institute for Health Research (NIHR) Clinical Lecturer Award (CL-2017-06-001, PIS), Health Education England Postdoctoral Research Fellowship (JME), Wellcome Trust Sir Henry Wellcome Fellowship (103986/Z/14/Z, CSM).

