## Supplementary materials for "North Carolina macular dystrophy: phenotypic variability and computational analysis of disease-implicated non-coding variants"

| **Supplementary Table 1:** Selected heterozygous single-nucleotide variants identified in the 560 kb region around the chr6:100,040,987G>C variant (corresponding to an identity-by-descent segment between families 75898 and 89794)* | | | | | | | | |
| --- | --- | --- | --- | --- | --- | --- | --- | --- |
| **chromosome** | **position**  (GRCh37) | **end** | **ref** | **alt** | **zygosity**  (case 75898.2) | **zygosity**  (case 89794.1) | **gnomAD allele count** | **gnomAD allele frequency** |
| chr6 | 100431554 | rs7773011 | T | C | het | het | 11027 | 0.352 |
| chr6 | 100431644 | rs7749425 | C | T | het | het | 11218 | 0.358 |
| chr6 | 100432655 | rs7754794 | C | T | het | het | 11387 | 0.364 |
| chr6 | 99948376 | rs75064329 | A | T | het | het | 193 | 0.006 |
| chr6 | 99956872 | rs72926237 | G | A | het | het | 1106 | 0.035 |
| chr6 | 99992605 | rs72926287 | G | A | het | het | 1036 | 0.040 |
| *Two study participants, 75898.2 and 89794.1, underwent genome sequencing. Sequencing data for patient 75898.2 were generated using the Complete Genomics platform and pipeline (v2.5) (see Ellingford *et al*.^14^ for further information). Sequencing in patient 89794.1 was performed by Edinburgh Genomics using the Illumina TruSeq Nano DNA kit and the Illumina MiSeq platform; a minimal median coverage of 30-fold was achieved and variants were assessed and filtered using the Variant Annotation and Filter Tool (VarAFT) (see de Bruijn *et al*.^28^ for further information).  After variant calling, genetic variation in the genomic region around the chr6:100,040,987G>C (GRCh37/hg19) change was evaluated in the two probands. The maximum presumed identity-by-descent segment around this mutation was then determined through the detection of discrepant homozygous calls between the probands; a 560 kb region, chr6:99867067-100433110 (GRCh37/hg19), was highlighted. Within this region we identified 219 variants that were both shared between the families and present in the Genome Aggregation Database (gnomAD). These included multiple heterozygous variants that had a minor allele frequency <5%; some of these changes are shown above. These observations suggest a shared rare haplotype (minor allele frequency <5%) flanking the chr6:100,040,987G>C variant between 75898.2 and 89794.1.  ref, reference sequence; alt, alternative sequence; gnomAD, Genome Aggregation Database v2.1.1; het, heterozygous. | | | | | | | | |

| **Supplementary Table 2:** Top 10 predicted *PRDM13*-linked enhancers detected using the Activity-by-Contact (ABC) method with chromatin accessibility data from human macular tissue as input | | | | | | |
| --- | --- | --- | --- | --- | --- | --- |
| **chr** | **start** | **end** | **activity** | **distance** | **HiC contact** | **ABC Score** |
| chr6 | 100036361 | 100036861 | 12.645519 | 18038 | 0.024038 | 0.010803 |
| chr6 | 100046330 | 100046830 | 3.361263 | 8069 | 0.056099 | 0.006515 |
| chr6 | 100038137 | 100039060 | 7.103467 | 16050 | 0.024038 | 0.006068 |
| chr6 | 100037142 | 100037642 | 6.886428 | 17257 | 0.024038 | 0.005883 |
| chr6 | 100049572 | 100050072 | 2.322015 | 4827 | 0.056099 | 0.004501 |
| chr6 | 100039139 | 100039639 | 3.774571 | 15260 | 0.024038 | 0.003225 |
| chr6 | 100040653 | 100041153 | 2.974766 | 13746 | 0.026677 | 0.002807 |
| chr6 | 100043983 | 100044483 | 2.138283 | 10416 | 0.026677 | 0.002017 |
| chr6 | 100066458 | 100066958 | 4.254949 | 12059 | 0.009265 | 0.00151 |
| chr6 | 100093765 | 100094265 | 2.213217 | 39366 | 0.013686 | 0.001116 |

| **Supplementary Table 3:** Top 10 predicted *PRDM13*-linked enhancers detected using the Activity-by-Contact (ABC) method with chromatin accessibility data from human retinal tissue as input | | | | | | |
| --- | --- | --- | --- | --- | --- | --- |
| **chr** | **start** | **end** | **activity** | **distance** | **HiC contact** | **ABC Score** |
| chr6 | 100036361 | 100036861 | 19.248011 | 18038 | 0.024038 | 0.018074 |
| chr6 | 100038619 | 100039588 | 14.615735 | 15545 | 0.024038 | 0.013724 |
| chr6 | 100046348 | 100046848 | 5.313465 | 8051 | 0.056099 | 0.01132 |
| chr6 | 100037098 | 100037598 | 9.831378 | 17301 | 0.024038 | 0.009232 |
| chr6 | 100093380 | 100094216 | 4.319864 | 39149 | 0.013686 | 0.002395 |
| chr6 | 100066419 | 100066919 | 4.759981 | 12020 | 0.009265 | 0.001856 |
| chr6 | 100091066 | 100091566 | 1.609914 | 36667 | 0.013686 | 0.000892 |
| chr6 | 100072214 | 100072714 | 2.321279 | 17815 | 0.007601 | 0.000762 |
| chr6 | 100027633 | 100028516 | 4.246895 | 26574 | 0.003256 | 0.000708 |
| chr6 | 100089821 | 100090321 | 1.099151 | 35422 | 0.013686 | 0.000609 |

| **Supplementary Table 3.** Expression levels of *PRDM13* as identified in single-cell RNA-seq studies of the human retina | | | | |
| --- | --- | --- | --- | --- |
| **Cell type** | **No. cells detected** | **Total cells** | **% of cells with detectable expression** | **Expression log2(CPM+1)** |
| Amacrine / horizontal cell precursors | 934 | 1891 | 49.39 | 5.23 |
| Amacrine cells | 10184 | 118913 | 8.56 | 0.88 |
| Horizontal cells | 2132 | 27388 | 7.78 | 0.79 |
| Neurogenic cells | 322 | 12978 | 2.48 | 0.25 |
| Photoreceptor precursors | 157 | 9887 | 1.59 | 0.16 |
| Pericytes | 77 | 9345 | 0.82 | 0.08 |
| Endothelial | 170 | 22138 | 0.77 | 0.07 |
| Retinal precursor cells | 285 | 37465 | 0.76 | 0.07 |
| Astrocytes | 23 | 3403 | 0.68 | 0.06 |
| Bipolar cells | 808 | 147915 | 0.55 | 0.06 |
| CPM, counts per million. All values were obtained from the <https://plae.nei.nih.gov/> resource. | | | | |

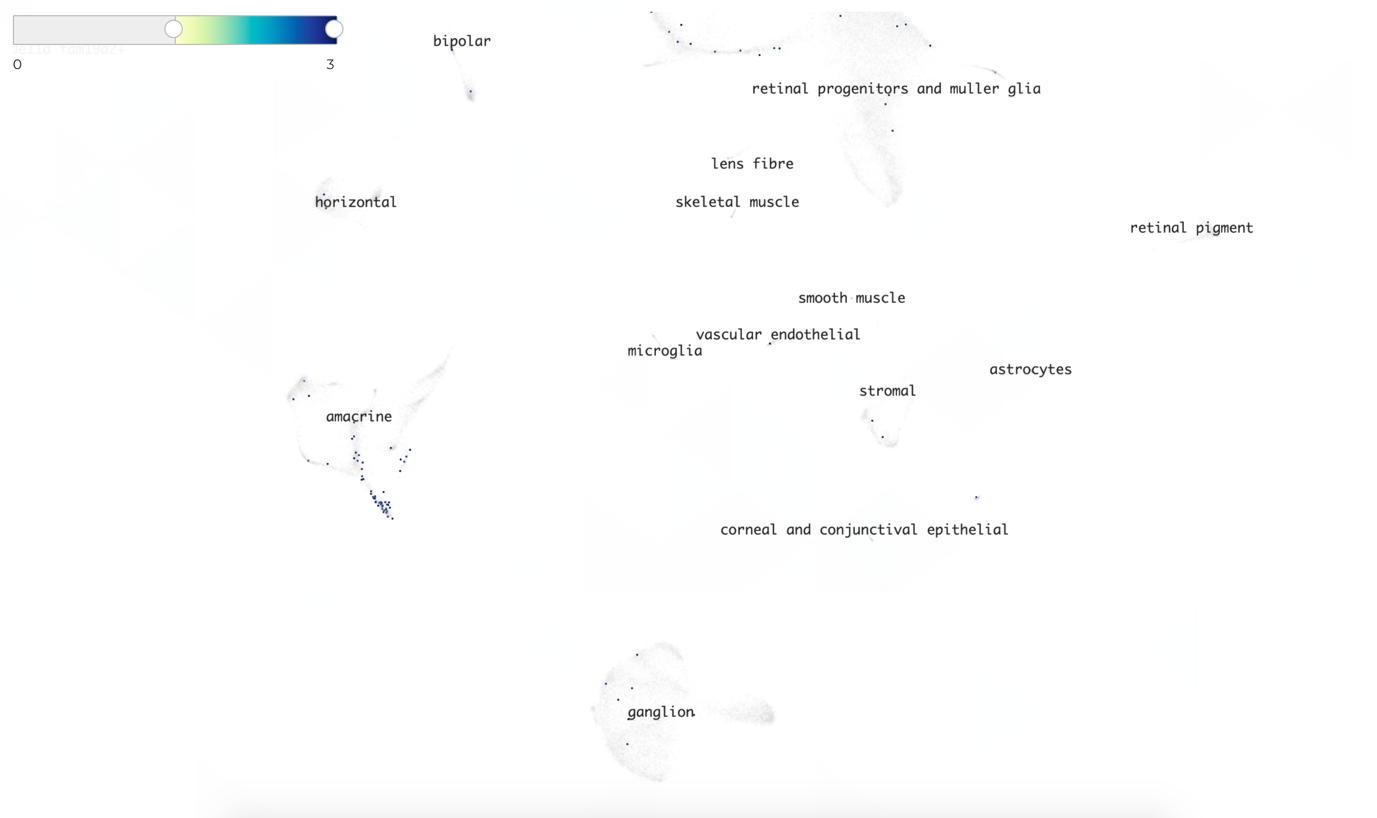

**Supplementary Figure 1:** UMAP (Uniform Manifold Approximation and Projection) plot showing single-cell expression of *PRDM13* in different cell types. Expression ranges from low (yellow) to high (blue). Cells with low expression have been hidden by setting the cut-off at 0.5. Data from <https://descartes.brotmanbaty.org/>.
